# Retrospect and prospect: a visual analysis of artificial intelligence applications in neurorehabilitation

**DOI:** 10.1101/2025.06.21.25330056

**Authors:** Tian Yuhan, Mai Tingting, Cai Mengcheng, Gu Wei, Fang Fanfu

## Abstract

**Objective:** To explore the current research status and future development trend of artificial intelligence in neurorehabilitation.

**Methods:** To collect and organize the literature on AI in neurorehabilitation from the core collection of Web of Science (WOS) database in the past ten years, and summarize the current status and predict the future development trend of AI in neurorehabilitation by using VOSviewer software and CiteSpace software in Java language environment.

**Results:** A total of 326 documents were retrieved, with the highest number of publications in 2014 (81); the scholars with the most publications were LiChong and PanYu (6); the institution with the most publications was Tsinghua University (8); the journal with the most publications was Sensors (26); and the number of Chinese research in the field of AI applied to neurorehabilitation is leading in the world. The keyword analysis shows that the research hotspots of AI applied to neurorehabilitation in recent years are mainly in brain-computer interface, deep learning, and robotics.

**Conclusion:** Recently, artificial intelligence in neurorehabilitation has developed rapidly, and the number of papers has increased year by year, mainly focusing on brain-computer interface, deep learning, and robotics.

## Introduction

Neurological Rehabilitation (NR) is a rehabilitation assessment and treatment for functional disorders such as motor, sensory, speech, swallowing, and cognitive disorders caused by neurological disorders, including stroke rehabilitation, cognitive rehabilitation, and motor rehabilitation. Stroke is the leading cause of disability globally, imposing a heavy socioeconomic burden while causing multiple impairments such as motor deficits, cognitive and emotional deficits ^[1]^, and there is a growing need for stroke rehabilitation, which is difficult to be met by existing means^**Error! Reference source not found**.^.

Artificial Intelligence (AI) is an emerging discipline that automates human intellectual tasks by simulating human intelligence behaviors and thought processes. And machine learning and deep learning are specific methods to achieve AI^[3]^. In recent years, although AI has been developing rapidly in the field of neurorehabilitation^[4]^, it is still in the early stage of development, and how to better apply AI in the field of neurorehabilitation is an issue that every neurorehabilitation healthcare practitioner needs to think about deeply^[5]^.

Bibliometrics is a tool to recognize the progress of a specific research field by means of analysis of vocabulary, citations, co-occurrence, etc. With the development of science and technology, the use of AI in the neurological rehabilitation field is getting hotter and hotter, and the rise of new AI technologies such as brain-computer interfaces has brought new therapeutic solutions for neurological rehabilitation. Analyzing the application of artificial intelligence in the neurorehabilitation field through bibliometrics is of great significance for researchers to explore more efficient and effective neurorehabilitation ^[6]^.

## Information and methodology

### Data sources

Data were obtained from the WOS Core Collection from January 1, 2015, to March 8, 2025, from the Science Citation Index Extension. The literature search strategy was as follows: ts=((“artificial intelligence” OR “AI” OR “machine learning” OR “deep learning” OR “neural network”) AND (“neurological rehabilitation” OR “neurorehab” OR “stroke rehabilitation” OR “brain injury rehabilitation” OR “cognitive rehabilitation” OR “motor recovery”)).

### Inclusion and exclusion criteria

Inclusion criteria: (1) compliance with the literature search strategy described above; (2) inclusion of treatises, reviews, and conference papers; (3) publication language designated as English. Exclusion criteria: (1) Literature and materials duplicated with the included literature; (2) Literature and materials not of thesis type such as scientific and technological achievements, books, etc.; (3) Literature and materials not related to the field of medical research, for example, literature involving animal husbandry, agriculture, and other fields.

### Research methodology

The research software included Java (version number 18), Microsoft Excel (version number 2019), and VOSviewer (version number 1.6.20). VOSviewer was run in a Java language environment to import the literature data provided by WOS Core Collection according to the desired format, and the visualization method was used to process the authors, issuing institutions, issuing countries, issuing journals, citation frequency of literature, and keywords.

## Results

### Analysis of trends in communications

Based on the inclusion and exclusion criteria, a total of 326 papers were included in this study. The frequency statistics of the number of literature publications showed that the number of articles published in 2015 was the lowest, 1; the number of articles published in 2024 was the highest, 81; and the number of articles showed an increasing trend from 2015 to 2024. The slope of the line graph shows that the number of publications of AI applied to NR grows slowly before 2019, and its number grows rapidly after 2019, see Figure 1: Annual Trends in Articles Number of articles/percentage of articles in vertical scale and year in horizontal scale. . Since the literature was acquired as of March 8, 2025, the number of publications in 2025 is not informative. In summary, the research heat of AI applied to the field of NR has gradually increased in the past decade, which may be due to the increasing attention on AI.

**Figure 1:**
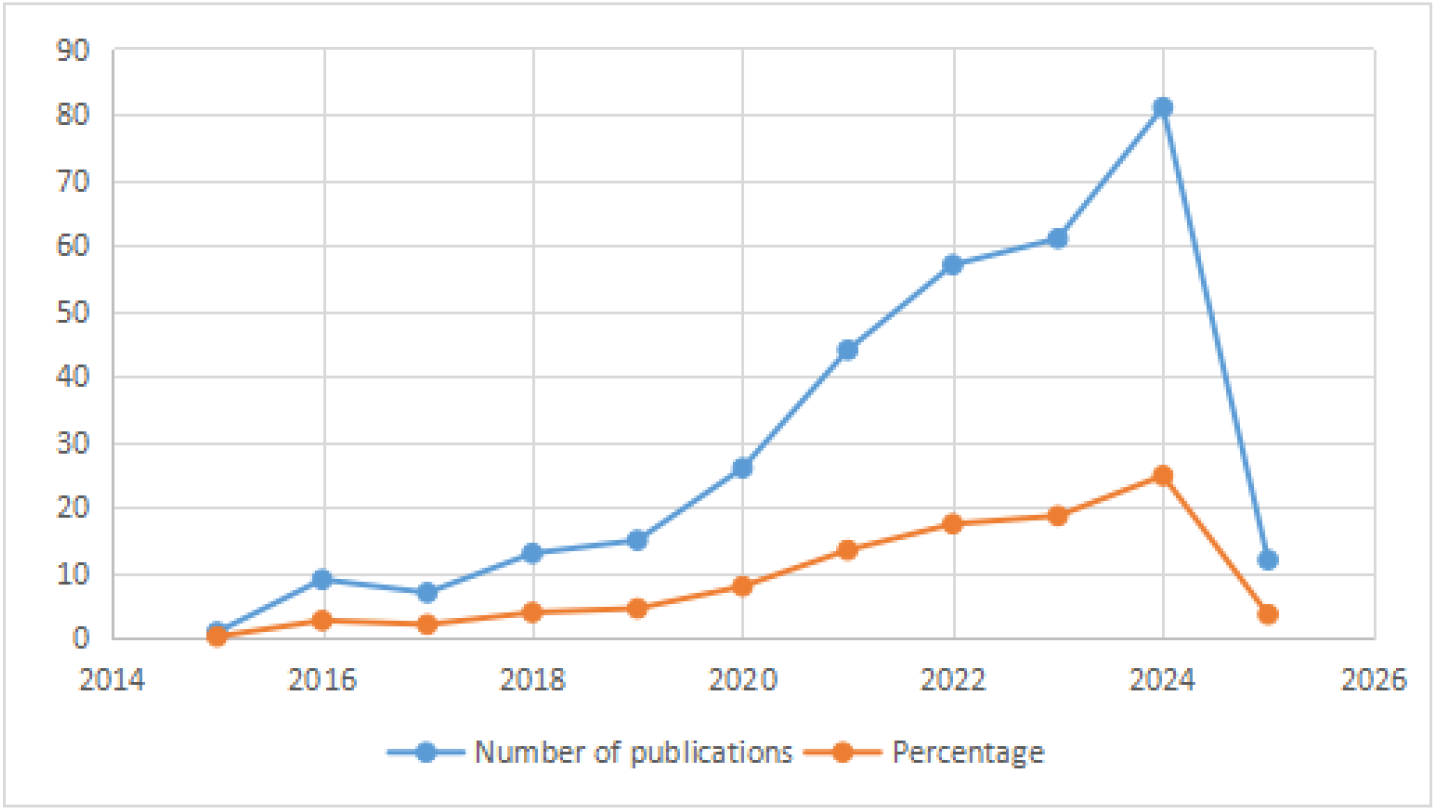
Annual Trends in Articles Number of articles/percentage of articles in vertical scale and year in horizontal scale.

### Analysis of journal article load and citation frequency

The top three journals with the most loaded articles are⟪Sensors⟫(26 articles), Neural System and Rehabilitation Engineering IEEE Transactions (16 articles), and Frontiers in Neuroscience (12 articles), and the results provide journal references for relevant researchers who want to study AI applied to NR (Table 1). Journal citation frequency can indicate the influence of the journal, to screen the journals with high influence, we analyzed the average citation frequency of AI applied to NR literature, and the results showed that the average citation frequency of the literature published by two journals, IEEE Transactions in Neurological System and Rehabilitation Engineering and Sensors, on the application of AI to the field of NR was higher, with 296 times and 277 times (Table 1), indicating that the two may be the core journals in the research of AI applied to the field of NR.

**Table 1:** Top 10 journals in terms of number of articles published.

| serial number | Journal Name | volume of publications | citation frequency |
| --- | --- | --- | --- |
| 1 | Sensors |  |  |
|  | Sensors | 26 | 296 |
| 2 | IEEE Transactions on Neurological and Rehabilitation Engineering. |  |  |
|  | Ieee transactions on neural systems and rehabilitation engineering | 16 | 277 |
| 3 | Frontiers in Neuroscience |  |  |
|  | Frontiers in neuroscience | 12 | 193 |
| 4 | The IEEE Interview |  |  |
|  | Ieee access | 11 | 172 |
| 5 | Journal of Neuroengineering and Rehabilitation |  |  |
|  | Journal of neuroengineering and rehabilitation | 11 | 150 |
| 6 | Frontiers in Neurology |  |  |
|  | Frontiers in Neurology | 9 | 131 |
| 7 | Biomedical Signal Processing and Control |  |  |
|  | Biomedical signal processing and control | 8 | 97 |
| 8 | Brain Science |  |  |
|  | Brain sciences | 7 | 82 |
| 9 | IEEE Sensors Magazine |  |  |
|  | Ieee sensors journal | 7 | 81 |
| 10 | Scientific Reports |  |  |
|  | Scientific reports | 7 | 35 |

### Analysis of issuing countries, issuing organizations, and authors

To understand which countries are the most prominent contributors to the field of AI and neurorehabilitation research, this study analyzed the publication volume of 52 countries. Figure 2 illustrates the visualization of country publication volume through VOSviewer. As can be seen in Figure 2, the distribution of countries publishing in this field is very uneven, and the top effect is significant. Further analyzing the highly productive countries in this field, Table 2 presents the top 5 countries in terms of publication volume in this field. As can be seen from Table 2, Chinese scholars contributed the most research papers in this field (a total of 102 publications), accounting for 31% of the total number of publications in this field. This is followed by the United States, with a total of 76 papers and 1,747 citations. The paper with the highest average number of citations was from Italy, with 24 papers obtaining 791 citations and an average number of citations as high as 33. The analysis results of the issuing institutions show (Figure 3): there are 22 institutions that published more than 4 (including 4) research articles related to the application of AI in the field of neurorehabilitation, of which the largest number of articles was issued by Tsinghua University (8), followed by the Chinese Academy of Sciences (7), Northwestern University (6), and Yonsei University (6).

**Figure 2:**
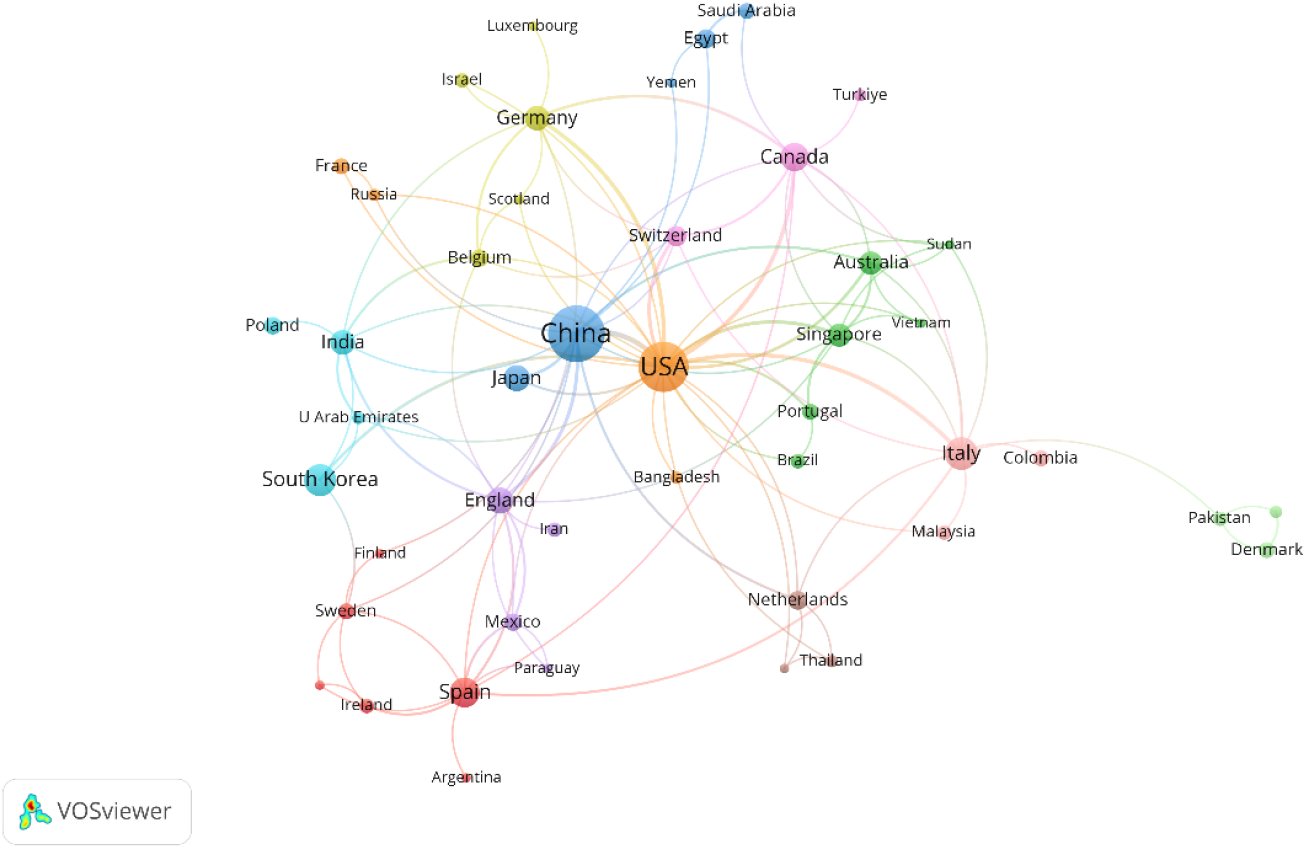
Co-occurrence map of issuing countries Note: The larger the round node, the more papers are sent: the node connecting line represents the strength of association, the thicker the line indicates that the two countries collaborate to send more papers; the node color represents different clusters.

**Figure 3:**
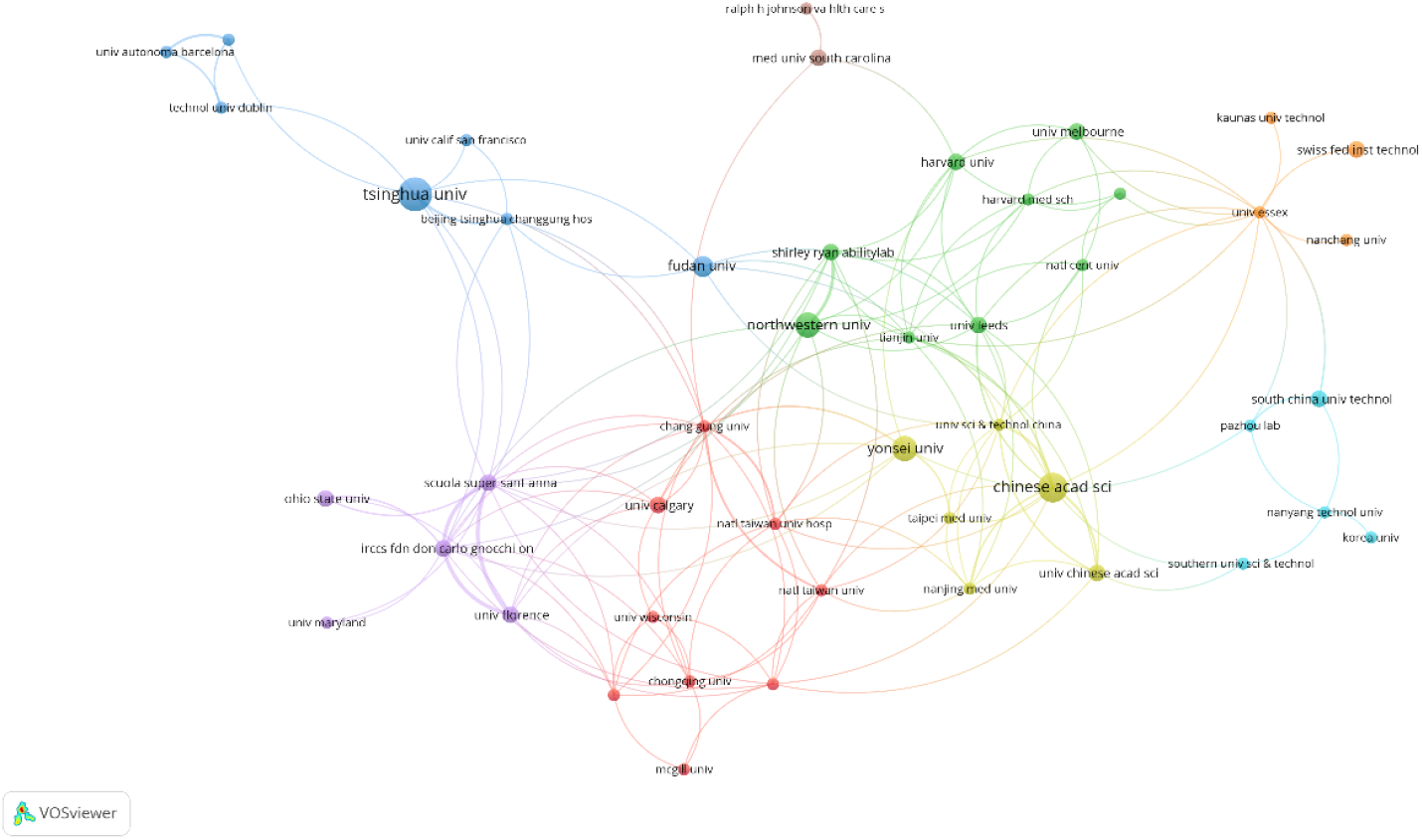
Co-occurrence map of issuing bodies Note: Larger node area means more documents are sent; more connectivity between nodes means higher intensity of cooperation.

**Table 2:** Top 5 countries in terms of the number of articles issued.

| Rank | Nations | Number of articles | Citations | Average number of citations |
| --- | --- | --- | --- | --- |
| 1 | China | 102 | 1145 | 11.2 |
| 2 | The United States | 76 | 1747 | 23.0 |
| 3 | Italy | 24 | 791 | 33.0 |
| 4 | South Korea | 23 | 337 | 14.7 |
| 5 | Spain | 19 | 168 | 8.8 |

Among the high-productivity authors, the most prolific publishers were LiChong of Tsinghua University and PanYu of Zhejiang University (Table 3). LiChong published a total of 6 papers from 2015 to March 2025, receiving 56 citations with an average number of citations of 9.3, and PanYu published 6 papers and received 62 citations, with an average number of citations of about 10.3 per paper.

**Table 3:**
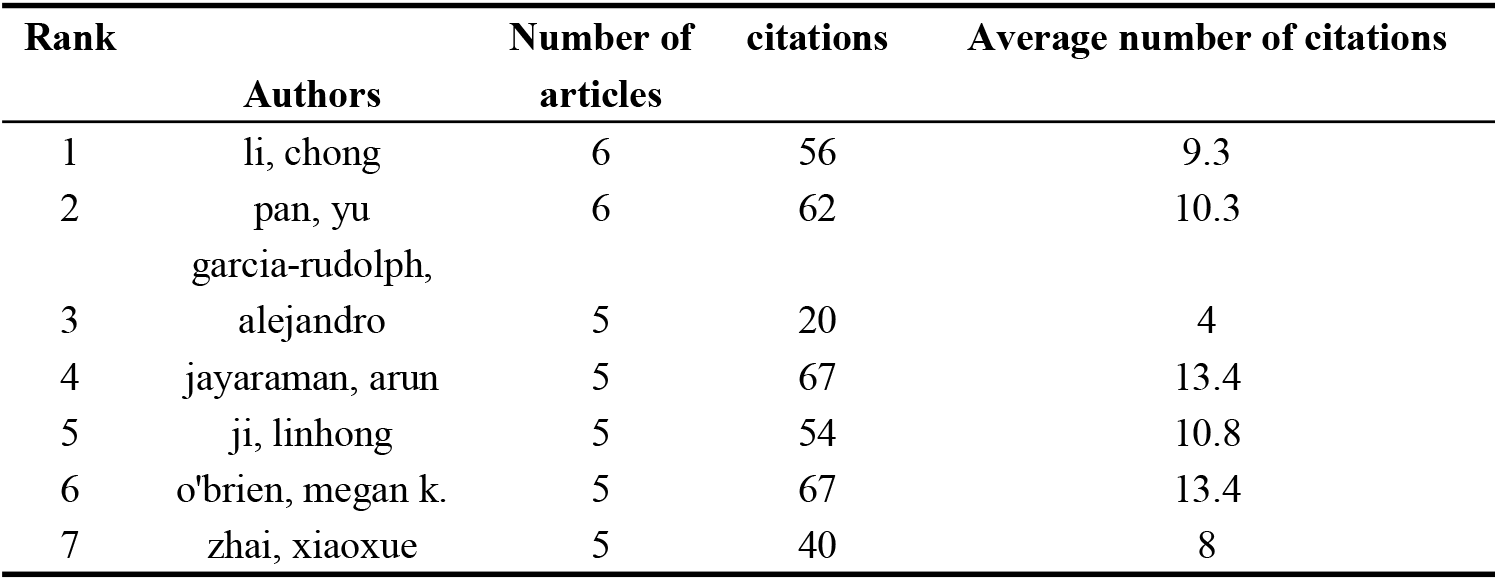
Top 7 authors in terms of number of articles published.

| Rank | Authors | Number of articles | citations | Average number of citations |
| --- | --- | --- | --- | --- |
| 1 | li, chong | 6 | 56 | 9.3 |
| 2 | pan, yu | 6 | 62 | 10.3 |
| 3 | garcia-rudolph, alejandro | 5 | 20 | 4 |
| 4 | jayaraman, arun | 5 | 67 | 13.4 |
| 5 | ji, linhong | 5 | 54 | 10.8 |
| 6 | o'brien, megan k. | 5 | 67 | 13.4 |
| 7 | zhai, xiaoxue | 5 | 40 | 8 |

### Frequency of literature citations

From the perspective of citation frequency (Table 4), Prof. Hamdi Altaheri’s paper “Deep learning techniques for classification of electroencephalogram (EEG) motor imagery (MI) signals: a review,” published in the journal Neural Computing and Applications in 2023 by Prof. Hamdi Altaheri has the highest citation frequency of 296. This study reviews the deep learning (DL)-based research on classification of electroencephalogram-motor imagery (MI-EEG) over the past decade, analyzes and discusses the DL-based techniques applied to MI classification from four main perspectives: preprocessing, input formulations, deep learning architectures, and performance evaluations, visualizes and analyzes the results, and discusses the challenges and directions of future development of AI technologies such as brain-computer interfaces (BCIs). challenges and directions^[7]^ . This literature has a relatively strong focus and influence on the research of AI applied to the field of NR.

**Table 4:** Top 10 cited articles.

| serial number | title | periodicals | citation frequency |
| --- | --- | --- | --- |
| 1 | Deep learning techniques for classification of electroencephalogram (EEG) motor imagery (MI) signals: a review | Neural Computing and Applications | 296 |
| 2 | Neurophysiological and Behavioral Effects of tDCS Combined With Constraint-Induced Movement Therapy in Poststroke Patients | Neurorehabilitation and Neural Repair | 240 |
| 3 | An end-to-end deep learning approach to MI-EEG signal classification for BCIs | Expert Systems with Applications | 234 |
| 4 | Daily Activity Monitoring and Outcome Assessments by Wearable Sensors | Neurorehabilitation and Neural Repair | 205 |
| 5 | EEG-Based Strategies to Detect Motor Imagery for Control and Rehabilitation | IEEE Transactions on Neural Systems and Rehabilitation Engineering | 170 |
| 6 | Action observation and motor imagery for rehabilitation in Parkinson's disease: a systematic review and an integrative hypothesis | Neuroscience & Biobehavioral Reviews | 139 |
| 7 | PP-Net: A Deep Learning Framework for PPG-Based Blood Pressure and Heart Rate Estimation | IEEE Sensors Journal | 130 |
| 8 | An IoT-Enabled Stroke Rehabilitation System Based on Smart Wearable Armband and Machine Learning | IEEE Journal of Translational Engineering in Health and Medicine | 120 |
| 9 | Repetitive transcranial magnetic stimulation at 1Hz and 5Hz produces sustained improvement in motor function and disability after ischaemic stroke. | European Journal of Neurology | 111 |
| 10 | Robotic Measurement of Arm Movements After Stroke Establishes Biomarkers of Motor Recovery | Stroke | 106 |

This is followed by Prof. Nadia Bolognini‘s paper “Neurophysiological and Behavioral Effects of tDCS Combined With Constraint-Induced Movement Therapy in Poststroke Patients”, published in 2011 in the journal Neurorehabilitation and Neural Repair, with 240 citations. Effects of tDCS Combined With Constraint-Induced Movement Therapy in Poststroke Patients” was published in the journal Neurorehabilitation and Neural Repair 240 times. In a double-blind controlled trial, it was found that traditional constraint-induced movement therapy could modulate local excitability but not transcallosal inhibition, while the addition of transcranial direct current stimulation could achieve greater functional recovery in both segments. It provides a practical and effective program for subsequent neurorehabilitation research^[8]^.

Prof. Hauke Dose’s paper “An end-to-end deep learning approach to MI-EEG signal classification for BCIs” was cited 234 times. The study demonstrates that a deep learning model using convolutional neural networks for the classification of motor intention EEG signals can be applied to the development of brain-computer interface systems, and that the model can be used as an alternative to established EEG classification methods by learning features from the data over time. This research lays the foundation for further development of brain-computer interfaces^[9]^.

Prof. Bruce H Dobkin’s 2011 publication in the journal Neurorehabilitation and Neural Repair, “Daily Activity Monitoring and Outcome Assessments by Wearable Sensors” published in the journal Neurorehabilitation and Neural Repair in 2011 was cited 205 times. The article analyzes current research and patient needs, predicts the rapid development of low-cost, energy-efficient wireless sensing and processing platforms for the future of healthcare delivery, clinical practice, and research, and offers practical recommendations for researchers^[10]^.

### Keyword co-occurrence analysis

Keywords condense the core and essence of a paper, and keyword co-occurrence analysis can be used to discover the research hotspots in a scientific field. Using VOSviewer to draw a keyword co-occurrence network view for 326 papers, Figure 4 visualizes the 125 keywords with a frequency greater than or equal to 5. To have a clearer understanding of the specifics of the keywords, the high-frequency keywords with a frequency of more than 6 were made into Table 5, from which it can be seen that the high-frequency keywords such as Motor recovery, Deep learning, and Machine learning constitute the representative terms in the field.

**Figure 4:**
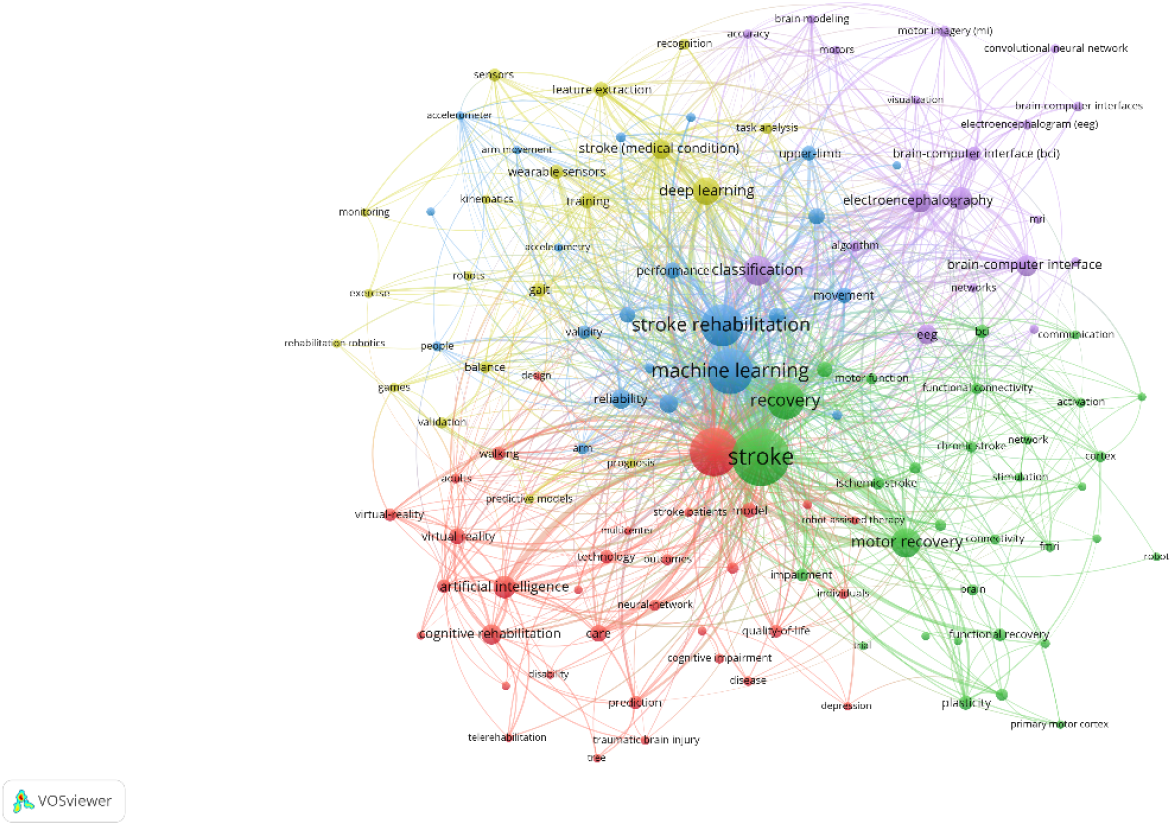
Keyword co-occurrence graph of literature. The larger area of the node represents the higher frequency of the word; the connecting lines between the nodes: the synergistic relationship between the keywords, the more lines, the higher the synergistic relationship.

**Table 5:** Top 10 keywords in terms of frequency of occurrence.

| Keyword | Occurrences | Total link strength |
| --- | --- | --- |
| Stroke | 116 | 191 |
| Rehabilitation | 89 | 172 |
| machine learning | 80 | 173 |
| stroke rehabilitation | 70 | 98 |
| recovery | 58 | 124 |
| motor recovery | 40 | 64 |
| classification | 39 | 75 |
| deep learning | 35 | 63 |
| artificial intelligence | 26 | 45 |
| Electroencephalography (EEG) | 25 | 53 |
| motor imagery | 25 | 46 |

From the evolution of keywords (Fig. 5), the time zone can be roughly divided into 2 stages: the research focus between 2015-2021 is relatively narrow, the combination of AI and neurorehabilitation is not deep enough, and the main focus is on some relatively shallow rehabilitation, such as pain inhibition, upper limb impact classification, etc.; between 2022-2025 there is an outbreak of research in the field, and brain-computer interfaces, Fugl-Meyer scale, predictive modeling, and other keyword clustering have all risen to prominence in this stage, becoming the mainstream of research, deep learning and other technologies have begun to enter the field in a big way, becoming the main method of research in the field, and began to pay attention to the rehabilitation system under the artificial intelligence science and technology, and the research as a whole has shown a tendency to go into the depths of several fields side by side.

**Figure 5:**
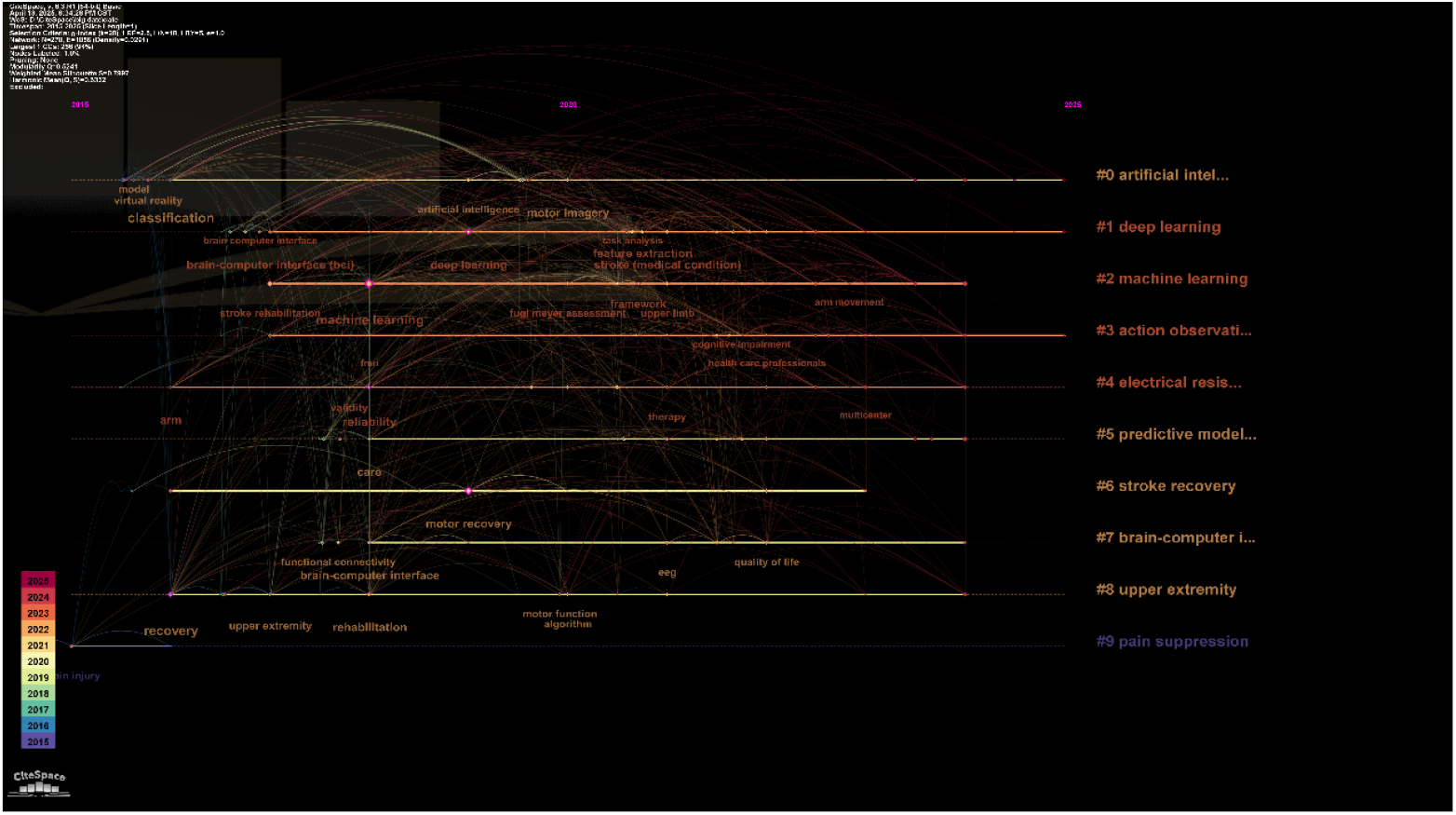
Keyword Time Zone Map. Horizontal coordinates are years, vertical coordinates are clusters.

Outbreak words can more clearly understand the sudden outbreak of research hotspots in the field of artificial intelligence and neurorehabilitation, outbreak word analysis function, the analysis results are shown in Figure 6. the results show that: the outbreak words are mostly concentrated in 2016-2023, and the research in this field during the period of 2020-2021 reflects the trend of the in-depth integration of rehabilitation and emerging technologies, and with the technology constantly being popularized and applied in the real rehabilitation treatment, the research focuses attention to brain function problems, and the emergence of thinking about the rehabilitation prognosis prediction model, in addition, descriptive statistics embodied in the trend of the issuance of articles can also be seen in the field after 2023, the growth of research in the field has reached its peak, and gradually tends to rationalization, the research direction is more mature.

**Figure 6:**
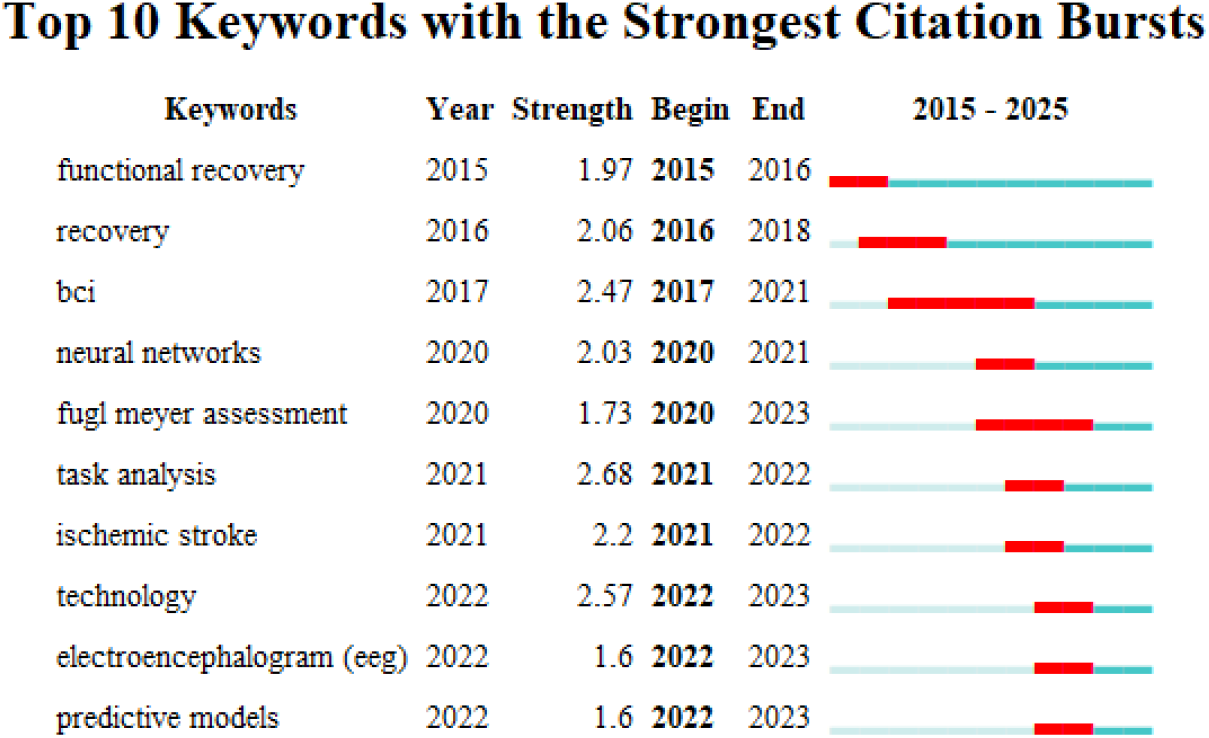
Top 10 burst word intensities

## Discussion

The WOS Core Collection was chosen as the data source for this study because it is a high-quality and comprehensive information resource retrieval database, and WOS is also considered to be the most suitable database for bibliometric analyses^[11]^ VOSviewer can show the hotspots of the research on the application of AI in the field of NR in a short period when performing keyword analysis; CiteSpace can explore the development history, current hotspots, and future trends of AI applied to NR by drawing time zone maps and outbreak word analysis maps. The combination of the two software complements each other to obtain more reliable, intuitive, and referential conclusions^[12]^. Therefore, based on VOSviewer and CiteSpace software, this study analyzes the research related to the application of AI to the field of NR in the past ten years of WOS, systematically reviews the development trend of the field, and analyzes the core authors, high-productivity countries, core journals in the field, and keyword clustering, which is of great significance for understanding and grasping the current status of its research and the current hotspots of the research.

Neurorehabilitation, as part of rehabilitation medicine, is an iterative field^[13]^. Artificial Intelligence is developing rapidly, and it is driving the healthcare system to high efficiency and a high level through big data analytics, etc^[14]^. Our study found that the trend of AI applied to the field of NR for research and published papers on it is on the rise^[15]^. The keyword analysis suggests that sports rehabilitation is one of the research hotspots in the field of AI applied to NR, and at the same time, sports rehabilitation is an important and already well-developed topic^[16,17]^.

Stroke-induced motor function deficits affect the patient’s mobility, resulting in limited activities of daily living, social participation, and reduced chances of returning to professional activities. The traditional process of motor rehabilitation treatment, accompanied by high-intensity, high-dose training and rehabilitation, has been slow to benefit from the effects^[18]^. And the combined use of non-invasive brain stimulation, robot-assisted training, and virtual reality immersion promises to achieve the maximum possible motor function recovery of patients^[19]^, which has a broad prospect in the future.

Keyword co-occurrence analysis also suggests that the brain-computer interface (BCI) is one of the research hotspots of PD to PF. In modern research, BCIs are widely used in stroke rehabilitation, where they translate brain signals and act as a neural signaling relay station between the paralyzed limb and the brain, and brain-computer interface combined with electrical stimulation therapies to promote functional recovery by activating the body’s natural efferent and afferent pathways^[20]^. Additionally, the potential for BCI to be used outside of the exercise domain is to improve cognitive and emotional rehabilitation in stroke patients, which may synergistically promote restorative neuroplasticity^[21]^. BCI has its advantages over traditional exercise rehabilitation, and is emerging as a new approach to post-stroke recovery^[22]^, but more clinical evidence is needed to confirm its clinical efficacy^[23]^.

Robotic exoskeleton is also one of the research hotspots suggested by keyword co-occurrence analysis. Currently, stroke-induced hemiparesis and arm paralysis are widely used in the rehabilitation process with passive assistive therapy devices and robotics in addition to exercise rehabilitation and BCI therapy^[24,25]^. Robotics for post-stroke rehabilitation is rapidly evolving, with a variety of robotic research hot off the press, with some studies suggesting the use of lightweight soft wearable robots, which provide relatively low levels of assistance sufficient to promote more normal ambulation in ambulatory individuals after stroke^[26]^. Many new randomized controlled trials have investigated the effects of robot-assisted treatment of upper limb paralysis, but have not failed to find a generalization of the improvement in upper limb ability, which may be related to a lack of understanding of robot-induced motor learning^[27]^, a field that is still under constant exploration and could be integrated with technologies such as BCI, with a promising future^[28]^.

There are some shortcomings in this study: this study only analyzed the literature on AI applications in NR where the language of publication was designated as English, and did not examine the literature in other languages. The scope of the literature has limitations, so future research needs to further expand the scope of data sources.

## Conclusion

This study uses bibliometrics to statistically and visually analyze the research overview of artificial intelligence in the field of neurorehabilitation. Currently, the research of artificial intelligence in the field of neurorehabilitation is in a period of steady development. At present, China has the highest number of publications in the world and has cooperated with other countries to a certain extent, indicating that China’s research in this field is in a more central leading position in the world. At present, the research heat of artificial intelligence in the field of neurorehabilitation is mainly focused on BCI, robotics, virtual reality, deep learning, etc. It is speculated that future research hotspots may focus on artificial intelligence and deep learning to convert neural signals.

## Data Availability

All relevant data are within the manuscript and its Supporting Information files.

## Acknowledgments

We would like to express our gratitude to those who provided help in data collecting and paper writing.

## Author Contributions

Conceptualization: Tian Yuhan,Mai Tingting.

Data curation: Tian Yuhan,Mai Tingting.

Formal analysis: Tian Yuhan.

Investigation: Fang Fanfu.

Methodology: Tian Yuhan,Mai Tingting,Cai Mengcheng.

Project administration: Gu Wei.

Software: Mai Tingting.

Supervision: Fang Fanfu.

Validation: Tian Yuhan,Mai Tingting,Cai Mengcheng.

Visualization: Tian Yuhan.

Writing – original draft: Tian Yuhan.

Writing – review & editing: Tian Yuhan,Mai Tingting.

